# Decomposing Interaction and Mediating Effects of Race/Ethnicity and Circulating Cystatin C on Cognitive Status in the United States Health and Retirement Study

**DOI:** 10.1101/2022.09.09.22279743

**Authors:** Cesar Higgins, Erin B. Ware, Lindsay C. Kobayashi, Mingzhou Fu, Margaret Hicken, Matthew Zawistowski, Bhramar Mukherjee, Kelly M. Bakulski

## Abstract

**Background and objectives:** Elevated circulating cystatin C is associated with cognitive impairment in non-Hispanic Whites, but its role in racial disparities in dementia is understudied. In a nationally representative sample of older non-Hispanic White, non-Hispanic Black, and Hispanic adults in the United States, we use mediation-interaction analysis to understand how racial disparities in the cystatin C physiological pathway may contribute to racial disparities in prevalent dementia.

**Methods:** In a pooled cross-sectional sample of the Health and Retirement Study (n=9,921), we employed Poisson regression to estimate prevalence ratios and to test the relationship between elevated cystatin C (>1.24mg/L versus <1.24mg/L) and impaired cognition, adjusted for demographics, behavioral risk factors, and other biomarkers. Self-reported racialized social categories were a proxy measure for exposure to racism. We calculated additive interaction measures and conducted four-way mediation-interaction decomposition analysis to test the moderating effect of race/ethnicity and mediating effect of cystatin C on the racial disparity.

**Results:** Overall, elevated cystatin C was associated with dementia (prevalence ratio [PR] = 1.4; 95%CI: 1.2, 1.8). Among non-Hispanic Black relative to non-Hispanic White participants, the relative excess risk due to interaction was 1.7 (95% CI: -0.2, 3.7), the attributable proportion was 0.2 (95%CI: 0.0, 0.5), and the synergy index was 1.4 (95% CI: 1.0, 2.0) in a fully-adjusted model. Elevated cystatin C was estimated to account for 2% (95% CI: -0%, 4%) for the racial disparity in prevalent dementia, and the interaction accounted for 9% (95% CI: -4%, 23%). Analyses for Hispanic relative to non-White participants suggested moderation by race/ethnicity, but not mediation.

**Discussion:** Elevated cystatin C was associated with dementia prevalence. Our mediation-interaction decomposition analysis suggested that the effect of elevated cystatin C on the racial disparity might be moderated by race/ethnicity, which indicates that the racialization process affects not only the distribution of circulating cystatin C across minoritized racial groups, but also the strength of association between the biomarker and dementia prevalence. These results provide evidence that cystatin C is associated with adverse brain health and this effect is larger than expected for individuals racialized as minorities had they been racialized and treated as non-Hispanic White.

## Introduction

Racial disparities in cognitive aging are well-documented,^1^ and thought to derive from exposure to structural and interpersonal forms of racism across the life course, including but not limited to residential and educational segregation,^2,3^ socioeconomic barriers,^4^ and psychosocial stress and discrimination.^5^ As a result, racism can affect diverse physiological pathways.^6^ Interpersonal forms of racism, such as perceived discrimination, have been associated with poor cognitive development^7^ and a moderate decline in kidney function^8^ among minoritized social groups. The link between chronic kidney disease and adverse cognitive outcomes has been documented through several studies^9–12^ and metanalyses.^13,14^ Racial minority groups in the United States are at increased risk for both adverse renal^15^ and cognitive outcomes.^1,16,17^ Understanding the share of cognitive impairment related to renal dysfunction across racial/ethnic groups may contribute to the identification of target areas for ameliorating racial disparities in cognitive outcomes.

Cystatin C is produced by all nucleated cells^18^ and its circulating blood levels are a marker of kidney function.^19,20^ Kidney function is related to brain health.^15^ In large population studies, circulating cystatin C varies by race/ethnicity, with higher blood levels observed in non-Hispanic Black compared to non-Hispanic White adults aged 50 to 60 years.^21^ However, prior studies of cystatin C and cognitive outcomes in older adults have not explored race-dependent relationships^9–12^—addressing this gap is essential for three main reasons. First, identifying subpopulations at most risk for dementia may provide insights for the highest impact interventions. Second, quantifying the mediating effect of cystatin C on the relationships between race/ethnicity and dementia may identify biological pathways underlying racial and ethnic disparities in cognition. Third, estimating the proportion of racial and ethnic disparities in dementia that could be reduced if cystatin C was amenable to intervention is useful to inform public health efforts and policies.

In a nationally representative racially/ethnically diverse sample of middle-aged and older adults in the United States, we tested the associations between circulating cystatin C levels and cognitive status. Using four-way mediation-interaction decomposition analysis,^22^ we aimed to examine the moderating effect of race/ethnicity in the association between cystatin C and cognitive status, and to understand how elevated levels of cystatin C may be a pathway (mediator) linking racial and ethnic disparities in cognitive impairment.

## Methods

### Study population

The United States Health and Retirement Study is an ongoing longitudinal study consisting of a representative sample of adults in the United States over age 50.^23^ To ensure representativeness of the United States population, recruitment is based on a multi-stage probability design with oversampling of underrepresented demographic groups in research (non-Hispanic Black and Hispanic participants). The initial cohort was formed in 1992 and interviews are conducted in 2-year waves.^24^ Primary respondents who meet age-eligibility criteria (age 50+) are randomly selected for interview along with a partner in the household, regardless of the partner’s age. For this analysis, we pooled a cross-sectional sample of participants who provided a blood spot at either the 2006 or 2008 wave. Proxy respondents were excluded, as they did not provide blood spots. Participants provided written informed consent at the time of the data collection. These secondary data analyses were approved by the University of Michigan Institutional Review Board (HUM00128220). Survey data are publicly available (https://hrs.isr.umich.edu/data-products), and genetic data are available through dbGaP (https://dbgap.ncbi.nlm.nih.gov; phs000428.v2.p2).

### Race and ethnicity

In the HRS, race is documented by self-reports across different waves of data. Study participants are asked to select their primary race from three large categories White/Caucasian, Black/African Americans, or Other. In an additional question, participants indicate whether they also identified as Hispanic or Latino. From these responses, we created a three mutually exclusive categorial variable (i.e., non-Hispanic Black, Hispanic, and non-Hispanic White categories). We used participants’ self-reported racialized category as a proxy indicator of exposure to racism and we compared cognitive status on each minoritized racial and ethnic category (non-Hispanic Black and Hispanic) to non-Hispanic White participants.

### Cystatin C

Dried blood spots were collected on a randomly selected half of the HRS cohort in the 2006 wave and the other half of the cohort in 2008. Levels of cystatin C were measured in blood spots using an optimized ELISA protocol.^25^ Assay performance was standardized to the European Certified Reference Material for cystatin C with Human Serum ERM-DA471/IFCC. Test results were verified by comparing 139 dried blood spot samples to matched plasma samples analyzed by microtiter plate ELISA. ELISA cystatin C levels were normalized to the United States National Health and Nutrition Examination Survey to ensure measures were comparable to venous blood and representative of the United States adult population. We considered quartiles of cystatin C in our exploratory analysis; and dichotomized cystatin C at the 75^th^ percentile (1.24 mg/L) in our regression models, this cutoff point was used in a previous research study.^9^ Participants with cystatin C levels greater than 1.24mg/L were considered high, and those less than or equal to 1.24mg/L were considered low.

To represent the additive interactions between elevated cystatin C levels and race/ethnicity, we operationalized cystatin C and each of race and ethnicity together (1 = non-Hispanic White participants with low cystatin C (unexposed group), 2 = non-Hispanic White participants with high cystatin C, 3 = non-Hispanic Black participants with low cystatin C, 4 = non-Hispanic Black participants with high cystatin C (double exposed)). We refer to the second and third levels as the “single-exposed” groups, because they have only one of the two exposures (minoritized race/ethnicity status or high cystatin C). We created a similar four-level variable for Hispanic participants, using non-Hispanic White participants with low cystatin C as the reference group. This operationalization allowed us to estimate the joint effect of race/ethnicity and cystatin C on cognitive function and directly estimate measures of interaction in the additive scale.^26^

### Cognitive assessment

Cognitive function was assessed by the Telephone Interview Cognitive Status, which provides a total score of 0 to 27 points based on a battery of cognitive tests assessing immediate and delayed recall of 10 words, serial 7s, counting backwards, object naming, and recalling the date, president, and vice-president to assess orientation.^27^ We used the Langa-Weir algorithm to classify cognitive function into three mutually exclusive categories: normal cognition (12 to 27), cognitive impairment non-dementia (CIND) (7 to 11), and dementia (0 to 6).^27^

### Covariate measures

Relevant covariates and known confounding factors between circulating cystatin C and cognitive status were identified from previous studies.^9–11^ Covariates were self-reported in the HRS interviews, and were: age (continuous, in years, calculated from birth date and interview date), sex (male; female), educational attainment (high school or less; college or some college; more than college), number of alcoholic drinks per day (continuous), smoking status (current smoker; former smoker; never smoker), body mass index (kg/meters^2^, continuous, based on self-reported height and weight), physician diagnosis of hypertension (Yes; No).

### Biomarkers

Other biomarkers measured in the dried blood spots were included as covariates.^25^ Levels of total cholesterol and high-density lipoprotein cholesterol were measured by microtiter plate assay using conventional clinical chemistry reactions optimized for the limited volume available from a dried blood spot sample. We constructed a ratio of total cholesterol to high density lipoprotein cholesterol as a marker of atherosclerosis. Levels of glycosylated hemoglobin (HbA1C) were measured on the dried blood spot; the assay was performed using Bio-Rad Laboratories Variant II High Pressure Liquid Chromatography (HPLC) System (Hercules, CA) optimized to accommodate the limited volume. We used HbA1C in is continuous form as a maker of glucose homeostasis.

### Genetic marker

Carriers of the Apolipoprotein E *ε4* allele (*APOE-*ε4) are at increased risk of cognitive impairment.^28,29^ Information on *APOE-ε4* allele carrier status was obtained from genetic data imputed to the worldwide 1,000 Genomes Project reference panel. Dosage data for rs7412 and rs429358 was used to determine *ε4* carrier status.^30^ We dichotomized the *APOE-ε4* carrier status into respondents with at least 1 copy of the allele *ε4* vs none and used carrier status in sensitivity models. Genotyping and imputation information on the Health and Retirement Study is available elsewhere.^31^

### Statistical analyses

We restricted our sample to participants with complete data for all covariates of interest. We described the bivariate relationships between quartiles of circulating cystatin C and categorical covariates using chi-square tests and with continuous covariates using ANOVA tests. We also described the distributions of covariates according to the combined cystatin C (high versus low) and race/ethnicity categories. Because cystatin C is a marker of renal function^32^ and kidney performance declines with age,^21^ we calculated age-group-specific (e.g., less than 60 years old, 60 to 79, 70 to 79, or 80 and more) predicted probabilities of high cystatin C (i.e., >1.24mg/L) for each racial/ethnic group. We plotted marginal predicted probabilities of high cystatin C (dependent variable) versus age (explanatory variable) to understand whether concentrations of elevated cystatin C varied across the three racialized group at different decades of life.

We used multivariable-adjusted Poisson regression models with a logarithmic link function and robust error variance^33^ to estimate prevalence ratios for the relationships between high cystatin C levels and each of CIND and dementia relative to normal cognition in the overall study sample and stratified by each racial group. We estimated four regression models: Model 1 was unadjusted, Model 2 was adjusted for demographic variables (age, sex, education), Model 3 was additionally adjusted for behavioral risk factors (smoking status, alcohol consumption, body mass index), and Model 4 was additionally adjusted for two biomarkers (the ratio of total cholesterol to high density lipoprotein cholesterol, HbA1C) and hypertension status. We calculated three measures of interaction in the additive scale, the relative excess risk due to interaction, the proportion attributable to the interaction, and the synergy index to evaluate effect modification of the association between cystatin C and prevalent dementia by racialized social groups.^26^ We performed four-way mediation-interaction decomposition analysis to evaluate whether cystatin C was a mediator of the racial disparity while allowing the mediator to interact with the exposure variable race/ethnicity.^22^ We illustrated the relationship between exposure, mediator, outcome, and confounding variables in a directed acyclic graph (**Supplemental Figure 1**). Detailed information on how interaction measures and the four-way mediation-interaction decomposition analysis were calculated can be found in the supplemental material.

### Sensitivity Analyses

*APOE-ε4* was not available for the full study sample. Thus, we tested *APOE-ε4* carrier status in a sensitivity analysis where the genetic marker was added to our previously specified demographic models (Model 2). These models were stratified by race/ethnicity. Because the Langa-Weir algorithm may have differential sensitivity across racial/ethnic groups, we also assessed dementia using the Power’s dementia algorithm (dementia versus normal cognition), a recently developed algorithm with comparable sensitivity of dementia classification across HRS racial groups.^34^

Analyses were conducted using R statistical software (version 3.6.2) and Stata (v14). For reproducibility purposes, we compared the code we developed to the Stata macro med4way.^35^ A second analyst performed complete code review. Code to produce these analyses is available (https://github.com/bakulskilab).

## Results

### Sample characteristics

There were 9,921 participants included in our analytic sample (**Supplemental Figure 2**). The sample had an average age of 68.4 years, with 12.6 years of education, and 1.12 mg/L circulating cystatin C levels (**Table 1**). In this sample, 13% of participants were non-Hispanic Black, 9% were Hispanic, 16% had cognitive impairment non-dementia, and approximately 4% had dementia. Excluded participants were slightly more likely to have CIND or dementia than included participants, but did not differ in their distributions of race, gender, education, alcohol consumption, smoking status, and hypertension (**Supplemental Table 1**). Excluded participants had similar concentrations of cystatin C to included participants, higher baseline HbA1C, and higher total cholesterol to high density lipoprotein cholesterol ratio.

**Table 1:** Sample characteristics by the percentiles of cystatin C distribution, United States Health and Retirement Study, 2006 and 2008

| Characteristic | Overall<br>N = 9,921<sup>1</sup> | cystatin C (mg/L) percentiles |  |  |  | p-value<sup>2</sup> |
| --- | --- | --- | --- | --- | --- | --- |
| | | 25<sup>th</sup><br>($\leq 0.86$)<br>N = 2,483<sup>1</sup> | 25<sup>th</sup>- 50<sup>th</sup><br>($> 0.86 \text{ \& } \leq 1.0$)<br>N = 2,601<sup>1</sup> | 50<sup>th</sup> - 75<sup>th</sup><br>($> 1.0 \text{ \& } \leq 1.24$)<br>N = 2,503<sup>1</sup> | 75<sup>th</sup><br>($> 1.24$)<br>N = 2,334<sup>1</sup> | |
| **Dementia status** |  |  |  |  |  | **<0.001** |
| Cognitively normal | 8,013 (96%) | 2,163 (98%) | 2,233 (98%) | 1,987 (95%) | 1,630 (91%) |  |
| Dementia | 346 (4.1%) | 42 (1.9%) | 52 (2.3%) | 95 (4.6%) | 157 (8.8%) |  |
| **Impairment status** |  |  |  |  |  | **<0.001** |
| Cognitively normal | 8,013 (84%) | 2,163 (89%) | 2,233 (88%) | 1,987 (83%) | 1,630 (75%) |  |
| Cognitively impaired non-dementia | 1,562 (16%) | 278 (11%) | 316 (12%) | 421 (17%) | 547 (25%) |  |
| **Race** |  |  |  |  |  | **<0.001** |
| Non-Hispanic Black | 1,270 (13%) | 339 (14%) | 309 (12%) | 284 (11%) | 338 (14%) |  |
| Hispanic | 918 (9.3%) | 296 (12%) | 249 (9.6%) | 208 (8.3%) | 165 (7.1%) |  |
| Non-Hispanic White | 7,733 (78%) | 1,848 (74%) | 2,043 (79%) | 2,011 (80%) | 1,831 (78%) |  |
| **Sex** |  |  |  |  |  | **<0.001** |
| Female | 5,957 (60%) | 1,595 (64%) | 1,530 (59%) | 1,450 (58%) | 1,382 (59%) |  |
| Male | 3,964 (40%) | 888 (36%) | 1,071 (41%) | 1,053 (42%) | 952 (41%) |  |
| **Education** |  |  |  |  |  | **<0.001** |
| > College | 889 (9.0%) | 307 (12%) | 256 (9.8%) | 181 (7.2%) | 145 (6.2%) |  |
| College/Some | 1,702 (17%) | 525 (21%) | 495 (19%) | 378 (15%) | 304 (13%) |  |
| High school or < | 7,330 (74%) | 1,651 (66%) | 1,850 (71%) | 1,944 (78%) | 1,885 (81%) |  |
| **Age (years)** | 68.37 (10.45) | 63.14 (9.26) | 66.47 (9.43) | 69.66 (9.68) | 74.68 (9.95) | **<0.001** |
| **Years of Education** | 12.58 (3.12) | 13.07 (3.12) | 12.80 (3.08) | 12.38 (3.03) | 12.02 (3.13) | **<0.001** |
| **Alcohol (# drinks/day when drinks)** | 0.69 (1.40) | 0.92 (1.49) | 0.80 (1.43) | 0.59 (1.25) | 0.42 (1.36) | **<0.001** |
| **Smoking Status** |  |  |  |  |  | **<0.001** |

| Characteristic | Overall | cystatin C (mg/L) percentiles |  |  |  | p-value<sup>2</sup> |
| --- | --- | --- | --- | --- | --- | --- |
|  |  | 25<sup>th</sup><br>(≤ 0.86)<br>N = 2,483<sup>1</sup> | 25<sup>th</sup>- 50<sup>th</sup><br>(> 0.86 & ≤ 1.0)<br>N = 2,601<sup>1</sup> | 50<sup>th</sup> - 75<sup>th</sup><br>(> 1.0 & ≤ 1.24)<br>N = 2,503<sup>1</sup> | 75<sup>th</sup><br>(> 1.24)<br>N = 2,334<sup>1</sup> |  |
| Current | 1,370 (14%) | 335 (13%) | 367 (14%) | 379 (15%) | 289 (12%) |  |
| Former | 4,284 (43%) | 994 (40%) | 1,138 (44%) | 1,079 (43%) | 1,073 (46%) |  |
| Never | 4,267 (43%) | 1,154 (46%) | 1,096 (42%) | 1,045 (42%) | 972 (42%) |  |
| **Body Mass Index (kg/m<sup>2</sup>)** | 28.30 (5.86) | 27.20 (5.33) | 28.15 (5.36) | 28.79 (5.93) | 29.10 (6.61) | **<0.001** |
| **Hypertension** |  |  |  |  |  | **<0.001** |
| No | 4,369 (44%) | 1,489 (60%) | 1,294 (50%) | 1,039 (42%) | 547 (23%) |  |
| Yes | 5,552 (56%) | 994 (40%) | 1,307 (50%) | 1,464 (58%) | 1,787 (77%) |  |
| **Baseline HbA1C (%)** | 5.86 (0.97) | 5.77 (1.03) | 5.79 (0.93) | 5.86 (0.91) | 6.03 (1.01) | **<0.001** |
| **Baseline TC/HDL-C (mg/dL)** | 3.97 (1.19) | 3.86 (1.16) | 3.92 (1.15) | 4.05 (1.24) | 4.04 (1.20) | **<0.001** |

<sup>1</sup>Statistics presented: N (Col%); Mean (SD)<sup>2</sup>Statistical tests performed: chi-square test of independence; One-way ANOVA
TC/HDL-C: total cholesterol to high density lipoprotein-cholesterol ratio; HbA1C: glycosylated hemoglobin

### Associations between cystatin C and cognitive status

Among those with cystatin C >1.24mg/L, the prevalence of CIND was 25%, and among those with levels <0.86mg/L was 11% (**Table 1**). Among those with cystatin C levels >1.24mg/L, the prevalence of dementia was 8.8%, and among those with levels <0.86mg/L was 1.9% (**Table 1 & Supplemental Figure 3**). Participants with cystatin C >1.24mg/L were more likely to be non-Hispanic White, to have high school education or less, and to be older than those with <0.86 mg/L cystatin C levels (**Table 1**). Among participants in their 50s through 70s, the non-Hispanic Black group had higher cystatin C than the other two racial/ethnic groups (**Figure 1**). Across all ages, the prevalence of high cystatin C was similar for the non-Hispanic White and Hispanic groups (**Figure 1**).

**Figure 1:**
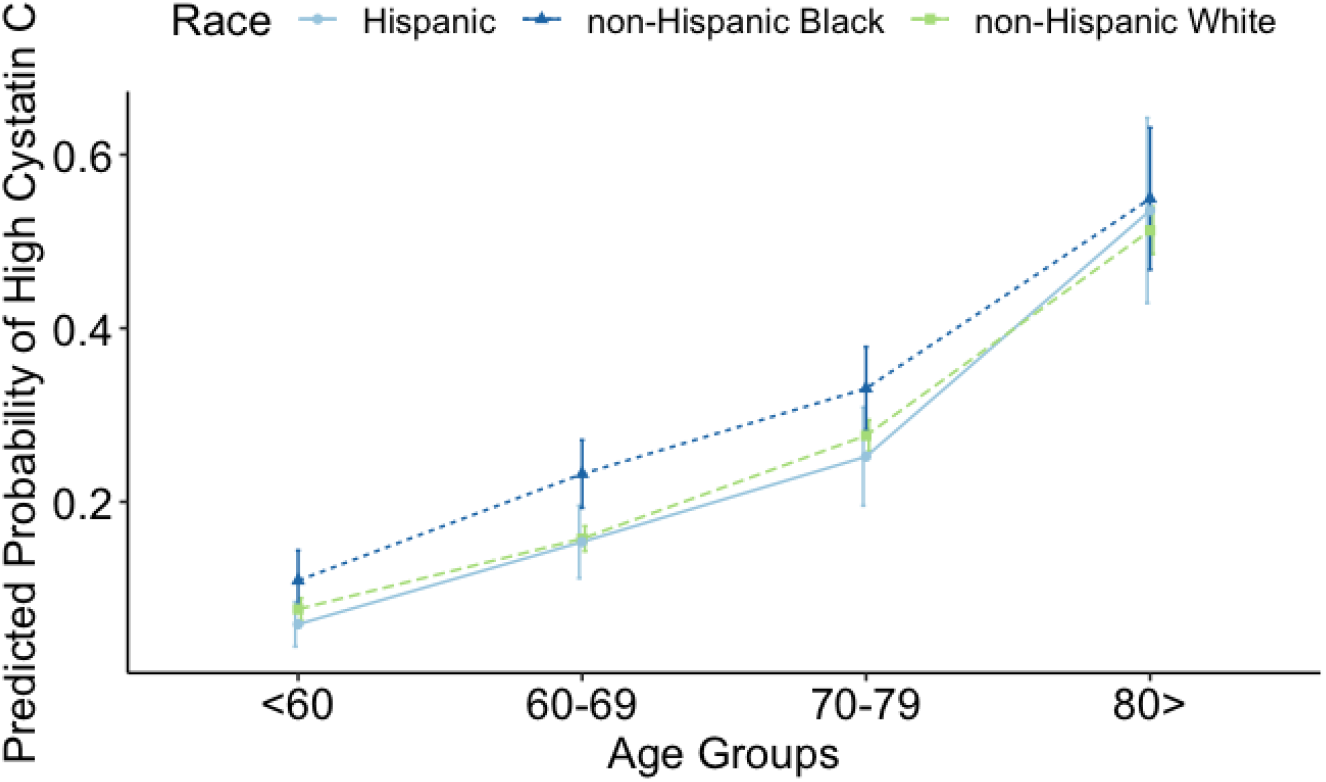
Marginal predicted probabilities of high cystatin C (>1.24mg/L) stratified by race/ethnicity in different age groups, in the Health and Retirement Study waves 2006 & 2008.

In our overall sample, model 4 showed that those with high cystatin C had 1.4 (95% CI: 1.2, 1.8) times higher prevalence of dementia and 1.2 (95% CI: 1.1, 1.3) times higher prevalence of CIND than those with low cystatin C levels (**Table 2**). The association between high cystatin C and dementia—but not cognitive impairment non-dementia—was similar across all racial groups (**Table 2**).

**Table 2:**
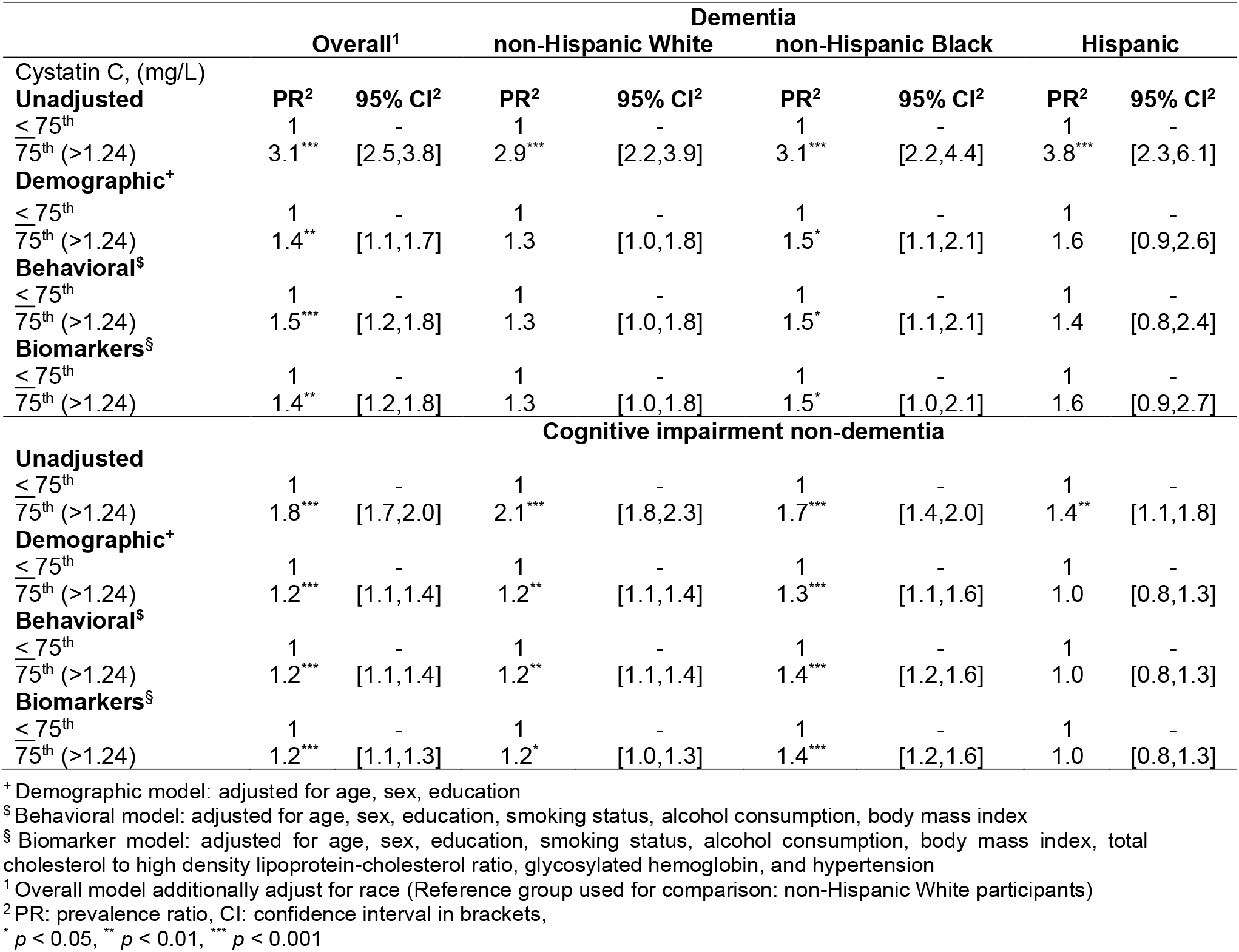
Adjusted prevalence ratios between high cystatin C levels (> 1.24mg/L) and dementia, and cognitive impairment non-dementia in the United States Health and Retirement Study stratified by race/ethnicity, waves 2006 & 2008

| Cystatin C, (mg/L) | Dementia |  |  |  |  |  |  |  |
| --- | --- | --- | --- | --- | --- | --- | --- | --- |
|  | Overall<sup>1</sup> |  | non-Hispanic White |  | non-Hispanic Black |  | Hispanic |  |
| **Unadjusted** | **PR<sup>2</sup>** | **95% CI<sup>2</sup>** | **PR<sup>2</sup>** | **95% CI<sup>2</sup>** | **PR<sup>2</sup>** | **95% CI<sup>2</sup>** | **PR<sup>2</sup>** | **95% CI<sup>2</sup>** |
| ≤ 75<sup>th</sup> | 1 | - | 1 | - | 1 | - | 1 | - |
| 75<sup>th</sup> (>1.24) | 3.1*** | [2.5,3.8] | 2.9*** | [2.2,3.9] | 3.1*** | [2.2,4.4] | 3.8*** | [2.3,6.1] |
| **Demographic<sup>+</sup>** |  |  |  |  |  |  |  |  |
| ≤ 75<sup>th</sup> | 1 | - | 1 | - | 1 | - | 1 | - |
| 75<sup>th</sup> (>1.24) | 1.4** | [1.1,1.7] | 1.3 | [1.0,1.8] | 1.5* | [1.1,2.1] | 1.6 | [0.9,2.6] |
| **Behavioral<sup>§</sup>** |  |  |  |  |  |  |  |  |
| ≤ 75<sup>th</sup> | 1 | - | 1 | - | 1 | - | 1 | - |
| 75<sup>th</sup> (>1.24) | 1.5*** | [1.2,1.8] | 1.3 | [1.0,1.8] | 1.5* | [1.1,2.1] | 1.4 | [0.8,2.4] |
| **Biomarkers<sup>§</sup>** |  |  |  |  |  |  |  |  |
| ≤ 75<sup>th</sup> | 1 | - | 1 | - | 1 | - | 1 | - |
| 75<sup>th</sup> (>1.24) | 1.4** | [1.2,1.8] | 1.3 | [1.0,1.8] | 1.5* | [1.0,2.1] | 1.6 | [0.9,2.7] |
| **Cognitive impairment non-dementia** |  |  |  |  |  |  |  |  |
| **Unadjusted** |  |  |  |  |  |  |  |  |
| ≤ 75<sup>th</sup> | 1 | - | 1 | - | 1 | - | 1 | - |
| 75<sup>th</sup> (>1.24) | 1.8*** | [1.7,2.0] | 2.1*** | [1.8,2.3] | 1.7*** | [1.4,2.0] | 1.4** | [1.1,1.8] |
| **Demographic<sup>+</sup>** |  |  |  |  |  |  |  |  |
| ≤ 75<sup>th</sup> | 1 | - | 1 | - | 1 | - | 1 | - |
| 75<sup>th</sup> (>1.24) | 1.2*** | [1.1,1.4] | 1.2** | [1.1,1.4] | 1.3*** | [1.1,1.6] | 1.0 | [0.8,1.3] |
| **Behavioral<sup>§</sup>** |  |  |  |  |  |  |  |  |
| ≤ 75<sup>th</sup> | 1 | - | 1 | - | 1 | - | 1 | - |
| 75<sup>th</sup> (>1.24) | 1.2*** | [1.1,1.4] | 1.2** | [1.1,1.4] | 1.4*** | [1.2,1.6] | 1.0 | [0.8,1.3] |
| **Biomarkers<sup>§</sup>** |  |  |  |  |  |  |  |  |
| ≤ 75<sup>th</sup> | 1 | - | 1 | - | 1 | - | 1 | - |
| 75<sup>th</sup> (>1.24) | 1.2*** | [1.1,1.3] | 1.2* | [1.0,1.3] | 1.4*** | [1.2,1.6] | 1.0 | [0.8,1.3] |

<sup>+</sup> Demographic model: adjusted for age, sex, education
<sup>§</sup> Behavioral model: adjusted for age, sex, education, smoking status, alcohol consumption, body mass index
<sup>§</sup> Biomarker model: adjusted for age, sex, education, smoking status, alcohol consumption, body mass index, total cholesterol to high density lipoprotein-cholesterol ratio, glycosylated hemoglobin, and hypertension
<sup>1</sup> Overall model additionally adjust for race (Reference group used for comparison: non-Hispanic White participants)
<sup>2</sup> PR: prevalence ratio, CI: confidence interval in brackets,
\* $p < 0.05$ , \*\* $p < 0.01$ , \*\*\* $p < 0.001$

### Joint associations between race/ethnicity and high cystatin C with cognitive status

Dementia was more prevalent among those who were non-Hispanic Black with high cystatin C (26% in this double exposed group) than those who were non-Hispanic Black with low cystatin C (8.2% in this single exposed group), non-Hispanic White with high cystatin C (5.5% in this single exposed group), or non-Hispanic White with low cystatin C (1.9% in this unexposed group; **Supplemental Table 2**). Similar patterns were observed for CIND. For Hispanic participants with high cystatin C levels (double exposed group), the prevalence of dementia and CIND was higher than Hispanic participants with low cystatin C (single exposed group), non-Hispanic White with high cystatin C (single exposed group), and non-Hispanic White with low cystatin C (unexposed group) (**Supplemental Table 3**).

In multivariable-adjusted models, we found that the double exposed group (non-Hispanic Black participants with high cystatin C) had higher prevalence of dementia than the unexposed group (non-Hispanic White participants with low cystatin C) (**Table 3**). Model 4 showed non-Hispanic Black participants with high cystatin C levels (double exposed group) had 7.0 (95% CI: 5.2, 9.4) times the prevalence of dementia than non-Hispanic White participants with low cystatin C (unexposed group) (**Table 3**). Similar findings were observed for Hispanic participants relative to non-Hispanic White participants. Model 4 showed that Hispanic participants with high cystatin C levels (double exposed group) had 5.8 (95%CI: 3.9, 8.7) times the prevalence of dementia than non-Hispanic White participants with low cystatin C (unexposed group) (**Table 3**). In general, for both minoritized racial groups, the associations between the double exposed compared to single exposed and unexposed groups in the CIND models were in the same direction but of lesser magnitudes to those observed in the dementia models (**Table 3**). However, in the Hispanic sample these associations were attenuated compared to the non-Hispanic Black sample.

**Table 3:** Prevalence ratios estimates from Poisson regression models illustrating the association between the joint effect of race/ethnicity and high cystatin C serum levels (> 1.24mg/L) and dementia or cognitively impaired non-dementia in the Health and Retirement Study, waves 2006 & 2008

| Models | Non-Hispanic Black |  |  |  |  |  | Hispanic |  |  |  |  |  |
| --- | --- | --- | --- | --- | --- | --- | --- | --- | --- | --- | --- | --- |
|  | Dementia |  |  | Cognitively impaired non-dementia |  |  | Dementia |  |  | Cognitively impaired non-dementia |  |  |
|  | PR<sup>1</sup> | 95% CI<sup>1</sup> | p-value | PR<sup>1</sup> | 95% CI<sup>1</sup> | p-value | PR<sup>1</sup> | 95% CI<sup>1</sup> | p-value | PR<sup>1</sup> | 95% CI<sup>1</sup> | p-value |
| **Unadjusted** |  |  |  |  |  |  |  |  |  |  |  |  |
| Unexposed<sup>a</sup> | 1 | — |  | 1 | — |  | 1 | — |  | 1 | — |  |
| Single exposed<sup>b</sup> (CysC) | 2.9 | 2.2,3.9 | <0.001 | 2.1 | 1.8,2.3 | <0.001 | 2.9 | 2.2,3.9 | <0.001 | 2.1 | 1.8,2.3 | <0.001 |
| Single exposed<sup>c</sup> (minority) | 4.4 | 3.2,6.0 | <0.001 | 2.9 | 2.6,3.3 | <0.001 | 3.1 | 2.1,4.6 | <0.001 | 2.7 | 2.3,3.1 | <0.001 |
| Double exposed<sup>d</sup> | 13.7 | 10.1,18.6 | <0.001 | 4.9 | 4.3,5.7 | <0.001 | 11.8 | 7.9,17.5 | <0.001 | 3.8 | 3.0,4.7 | <0.001 |
| **Demographic<sup>+</sup>** |  |  |  |  |  |  |  |  |  |  |  |  |
| Unexposed<sup>a</sup> | 1 | — |  | 1 | — |  | 1 | — |  | 1 | — |  |
| Single exposed<sup>b</sup> (CysC) | 1.4 | 1.0,1.8 | <0.05 | 1.3 | 1.1,1.5 | <0.001 | 1.3 | 1.0,1.8 | >0.05 | 1.3 | 1.1,1.5 | <0.001 |
| Single exposed<sup>c</sup> (minority) | 5.0 | 3.7,6.8 | <0.001 | 3.0 | 2.7,3.4 | <0.001 | 3.9 | 2.7,5.6 | <0.001 | 2.9 | 2.5,3.3 | <0.001 |
| Double exposed<sup>d</sup> | 7.2 | 5.4,9.6 | <0.001 | 3.7 | 3.2,4.2 | <0.001 | 5.8 | 3.9,8.6 | <0.001 | 2.5 | 2.0,3.1 | <0.001 |
| **Behavioral<sup>§</sup>** |  |  |  |  |  |  |  |  |  |  |  |  |
| Unexposed<sup>a</sup> | 1 | — |  | 1 | — |  | 1 | — |  | 1 | — |  |
| Single exposed<sup>b</sup> (CysC) | 1.4 | 1.0,1.9 | <0.05 | 1.3 | 1.1,1.5 | <0.001 | 1.4 | 1.0,1.8 | >0.05 | 1.3 | 1.1,1.4 | <0.001 |
| Single exposed<sup>c</sup> (minority) | 5.2 | 3.8,7.1 | <0.001 | 3.1 | 2.7,3.5 | <0.001 | 3.9 | 2.7,5.7 | <0.001 | 2.9 | 2.5,3.3 | <0.001 |
| Double exposed<sup>d</sup> | 7.4 | 5.5,9.9 | <0.001 | 3.7 | 3.2,4.3 | <0.001 | 6.1 | 4.1,9.2 | <0.001 | 2.5 | 2.0,3.1 | <0.001 |
| **Biomarkers<sup>§</sup>** |  |  |  |  |  |  |  |  |  |  |  |  |
| Unexposed<sup>a</sup> | 1 | — |  | 1 | — |  | 1 | — |  | 1 | — |  |
| Single exposed<sup>b</sup> (CysC) | 1.3 | 1.0,1.8 | >0.05 | 1.3 | 1.1,1.4 | <0.001 | 1.3 | 1.0,1.8 | >0.05 | 1.2 | 1.1,1.4 | <0.01 |
| Single exposed<sup>c</sup> (minority) | 4.9 | 3.6,6.7 | <0.001 | 2.9 | 2.6,3.3 | <0.001 | 3.5 | 2.4,5.1 | <0.001 | 2.8 | 2.4,3.2 | <0.001 |
| Double exposed<sup>d</sup> | 7.0 | 5.2,9.4 | <0.001 | 3.5 | 3.0,4.1 | <0.001 | 5.8 | 3.9,8.7 | <0.001 | 2.4 | 1.9,3.0 | <0.001 |
| **Observations** | 7,683 |  |  | 8,715 |  |  | 7,481 |  |  | 8,412 |  |  |

<sup>1</sup> PR: prevalence ratio, CI: confidence interval
CysC: Cystatin C
<sup>a)</sup> Unexposed group (reference group): non-Hispanic White & Low cystatin C ( $\leq 1.24\text{mg/L}$ )
<sup>b)</sup> Single exposed (CysC): non-Hispanic White & High cystatin C ( $> 1.24\text{mg/L}$ )
- c) Single exposed (minority): either non-Hispanic Black or Hispanic & Low cystatin C ( $\leq 1.24\text{mg/L}$ ) - d) Double exposed: either non-Hispanic Black or Hispanic & High cystatin C ( $> 1.24\text{mg/L}$ ) - + Demographic model: adjusted for age, sex, education - \$ Behavioral model: adjusted for age, sex, education, smoking status, alcohol consumption, body mass index - § Biomarker model: adjusted for age, sex, education, smoking status, alcohol consumption, body mass index, total cholesterol to high density lipoprotein-cholesterol ratio, glycosylated hemoglobin, and hypertension

### Measures of interaction on the additive scale between race/ethnicity, cystatin C, and dementia prevalence

Among non-Hispanic Black participants, model 4 yielded a relative excess risk due to interaction of 1.7 (95% CI: -0.2, 3.7), indicating that the joint effect of having a double exposure exceeded the simple addition of being either a non-Hispanic Black participant or having high cystatin C (**Table 4**). Additionally, from the same model 4 we estimated an attributable proportion of 0.2 (95% CI: 0.0, 0.5), which suggests that 20% of the relative excess risk of dementia in the double exposed group may be attributable to the interaction between the two main exposures (race/ethnicity and high cystatin C). The synergy index represents the quotient between the prevalence ratio of dementia in the double exposed group divided by the isolated prevalence ratios of each single exposed group, a quotient greater than one indicates possible synergistic effect or effect modification. In model 4, he synergy index was 1.4 (95% CI: 1.0, 2.0), indicating synergism between both exposures. We obtained similar estimates for the other models.

**Table 4:** Measures of interaction in the additive scale of the joint effect of race/ethnicity and high cystatin C (>1.24mg/L) on dementia. Additive measures calculated using prevalence ratios from Poisson regression models (in Table 3) stratified by race/ethnicity.

| Models | Non-Hispanic Black<sup>a</sup> |  |  | Hispanic<sup>a</sup> |  |  |
| --- | --- | --- | --- | --- | --- | --- |
|  | Estimate | 95% CI<sup>1</sup> | p-value | Estimate | 95% CI<sup>1</sup> | p-value |
| **Unadjusted** |  |  |  |  |  |  |
| Excess risk due to interaction<sup>†</sup> | 7.4 | 3.7, 11.1 | **<0.001** | 6.7 | 2.3, 11.1 | **<0.01** |
| Attributable proportion<sup>‡</sup> | 0.5 | 0.4, 0.7 | **<0.001** | 0.6 | 0.4, 0.8 | **<0.001** |
| Synergy index<sup>£</sup> | 2.4 | 1.7, 3.3 | **<0.001** | 2.7 | 1.7, 4.5 | **<0.001** |
| **Demographic<sup>+</sup>** |  |  |  |  |  |  |
| Excess risk due to interaction<sup>†</sup> | 1.8 | -0.1, 3.8 | 0.07 | 1.6 | -0.8, 3.9 | 0.19 |
| Attributable proportion<sup>‡</sup> | 0.3 | 0.0, 0.5 | **0.03** | 0.3 | -0.1, 0.6 | 0.12 |
| Synergy index<sup>£</sup> | 1.4 | 1.0, 2.0 | 0.07 | 1.5 | 0.8, 2.7 | 0.17 |
| **Behavioral<sup>§</sup>** |  |  |  |  |  |  |
| Excess risk due to interaction<sup>†</sup> | 1.8 | -0.2, 3.8 | 0.09 | 1.8 | -0.7, 4.3 | 0.15 |
| Attributable proportion<sup>‡</sup> | 0.2 | 0.0, 0.5 | **0.05** | 0.3 | -0.0, 0.6 | 0.07 |
| Synergy index<sup>£</sup> | 1.4 | 1.0, 2.0 | 0.09 | 1.5 | 0.9, 2.7 | 0.13 |
| **Biomarkers<sup>§</sup>** |  |  |  |  |  |  |
| Excess risk due to interaction<sup>†</sup> | 1.7 | -0.2, 3.7 | 0.08 | 2.0 | -0.4, 4.3 | 0.10 |
| Attributable proportion<sup>‡</sup> | 0.2 | 0.0, 0.5 | **0.04** | 0.3 | 0.2, 0.6 | **0.03** |
| Synergy index<sup>£</sup> | 1.4 | 1.0, 2.0 | 0.08 | 1.7 | 0.9, 3.0 | 0.08 |
| **Observations** | 7,683 |  |  | 7,481 |  |  |

<sup>a</sup>) Reference group used for comparison: non-Hispanic White participants
<sup>+</sup> Demographic model: adjusted for age, sex, education
<sup>§</sup> Behavioral model: adjusted for age, sex, education, smoking status, alcohol consumption, body mass index
<sup>§</sup> Biomarker model: adjusted for age, sex, education, smoking status, alcohol consumption, body mass index, total cholesterol to high density lipoprotein-cholesterol ratio, glycosylated hemoglobin, and hypertension
<sup>†</sup> Excess risk = Prevalence ratio (PR) of double exposed - PR of single exposed (minority) - PR of single exposed (CysC) + 1
<sup>‡</sup> Attributable proportion = [Excess risk due to interaction] / [PR of double exposed]
<sup>£</sup> Synergy index = [PR of double exposed - 1] / [PR of single exposed (minority) + PR of single exposed (CysC) - 2]
<sup>1</sup> CI: confidence interval calculated using the delta method

Point measures of additive interaction for the Hispanic group were equivalent with slightly larger confidence intervals to those observed for non-Hispanic Black participants (**Table 4**). Estimates for both groups suggest a possible interaction effect between minoritized race/ethnicity status and high cystatin C. Although some point estimates were not statistically significant, all measures of additive interaction were positive and suggested synergism. We did not observe additive interaction between race/ethnicity and high cystatin C on CIND prevalence.

### Four-way mediation-interaction decomposition to assess race/ethnicity and cystatin C on dementia prevalence

The decomposition analysis comparing non-Hispanic Black to White participants, adjusting for demographic variables, showed that the mediating effect of cystatin C accounted for 2% (95% CI: 0%, 4%) of the observed racial disparity in dementia (**Table 5**), while the portion attributable to the interaction between exposure and mediator (moderating pathway) accounted for 9% (95% CI: -4%, 23%) of the disparity. The decomposition analysis indicated that 90% (95% CI: 77%, 104%) of the racial disparity was due to the controlled direct effect of race/ethnicity, pointing that, of course, there are other pathways through which racism operates besides high cystatin C levels (**Table 5**). These attributable proportions can be interpreted causally if a sufficient set of confounder variables have been identified and no violations of the causal mediation assumptions are present.^22^

**Table 5:** Decomposition estimates of mediation analysis results, race/ethnicity used as proxy exposure for racism and high serum cystatin C levels (>1.24mg/L) as mediator. Models allow for exposure-mediator interaction effects. Estimates presented by race/ethnic groups

| **Mediation-Interaction Decomposition<sup>†</sup>** |  |  |  |  |  |  |  |  |
| --- | --- | --- | --- | --- | --- | --- | --- | --- |
| **Component** | **non-Hispanic Black<sup>a</sup>** |  |  |  | **Hispanic<sup>a</sup>** |  |  |  |
|  | **Excess**<br>**Relative**<br>**Risk** | **95% CI** | **Proportion**<br>**Attributable** | **95% CI** | **Excess**<br>**Relative**<br>**Risk** | **95% CI** | **Proportion**<br>**Attributable** | **95% CI** |
| Controlled Direct Effect | 4.00 | 2.4, 5.6 | 90.0% | 77%, 104% | 2.90 | 1.34, 4.46 | 91.0% | 75%, 107% |
| Interaction Reference | 0.35 | -0.1, 0.8 | 8% | -3%, 19% | 0.30 | -0.21, 0.82 | 9.0% | -8%, 27% |
| Interaction Mediation | 0.07 | -0.0, 0.2 | 2% | -1%, 4% | -0.02 | -0.04, 0.01 | 0% | -1%, 0% |
| Pure Indirect Effect | 0.01 | -0.0, 0.0 | 0.0% | 0%, 1% | -0.00 | -0.01, 0.00 | 0% | 0%, 0% |
| Total Effect | 4.43 | 2.9, 5.9 | 100% |  | 3.18 | 1.76, 4.60 | 100% |  |
| **Overall Proportion** |  |  |  |  |  |  |  |  |
| Due to Mediation |  |  | 2% | -0%, 4% |  |  | -1% | -1%, 0% |
| Due to Interaction |  |  | 9% | -4%, 23% |  |  | 9% | -7%, 25% |

<sup>a)</sup> Reference group used for comparison: non-Hispanic White participants
<sup>†</sup> Demographic model: adjusted for age, sex, education

For the Hispanic group, the demographic-adjusted decomposition analysis showed similar interaction results to that of the non-Hispanic Black group. Among Hispanic participants, the mediating effect of cystatin C in the racial disparity was virtually zero, while the portion attributable to the interaction accounted for 9% (95% CI: -7%, 25%) of the racial disparity and the direct effect for 91% (95% CI: 75%, 107%) (**Table 5**). We did not estimate attributable fractions for models controlling for behavioral risk factors, or other biomarkers, as the mediating effect of cystatin C was negligible for both minoritized racial groups with further adjustment.

### Sensitivity Analyses

We re-estimated measures of additive interaction to test whether the omission of the genetic influence of *APOE-ε4* on dementia biased our mediation decomposition results. After adjusting for *APOE-ε4*, prevalence ratios of dementia for the double and single exposed groups (**Supplemental Table 4**) were consistent to our main fully adjusted models without the inclusion of *APOE-ε4* (**Table 4**).

To assess robustness of our findings to potential dementia misclassification, we repeated our analyses using Power’s two-level dementia outcome. In the demographic-adjusted model, among non-Hispanic Black participants with high cystatin C (double exposed group), the prevalence of dementia was 2.6 (95%CI: 2.0, 3.4) times higher than for non-Hispanic White participants with low cystatin C (unexposed group) (**Supplemental Table 5A**). For Hispanic participants, using Power’s algorithm did not show an association between Hispanic ethnicity, high cystatin C levels, and dementia (**Supplemental Table 5B**). Additionally, measures of interaction in the additive scale were close to the null value and non-significant for both minoritized social groups (**Supplemental Table 6**).

## Discussion

In this large and diverse cross-sectional analysis of older adults in the United States, high levels of serum cystatin C (>1.24 mg/L) were associated with elevated prevalence of cognitive impairment non-dementia and dementia relative to normal cognition. The association between high cystatin C and dementia was suggestive of super-additivity for minoritized racial groups, but the confidence intervals for the additive measures of interaction were imprecise. We found that 9% of the disparity in prevalent dementia between non-Hispanic Black and non-Hispanic White participants was due to the interaction between race and high cystatin C levels, while 2% was mediated through cystatin C. Altogether, these results suggest that, among older adults who are racialized as non-Hispanic Black or Hispanic, the effect of high cystatin C levels on dementia prevalence is greater than expected had these individuals been racialized and treated as non-Hispanic White.^3637,38^ However, the largest proportion of the observed disparity could be thus attributable to other pathways through which racism operates besides cystatin C.^36^

Three observational studies previously documented an association between elevated cystatin C levels and adverse cognitive health outcomes.^9–11^ The Aging Brain Cohort (ABC) study, another large racially diverse study conducted in the United States, found that individuals with cystatin C serum levels >1.25mg/L had 1.92 times higher odds of 7-year incident cognitive impairment with respect to those with cystatin C levels <1.0mg/L.^9^ The ABC study limited race-dependent analysis to a multiplicative interaction; however, we expanded on their analysis by testing additive interactions between cystatin C and race/ethnicity. The Cardiovascular Health Study, a community-based longitudinal cohort of older adults (aged >65) with a representative sample of non-Hispanic Black participants, estimated a 1.39 greater hazard of cognitive impairment among those with cystatin C levels >1.16mg/L vs. <0.90mg/L.^11^ This study did not explore an interaction between race and cystatin C. Lastly, a prospective cohort of women aged >65 in the United States showed that cystatin C levels of 1.15-2.37mg/L (vs. 0.61-0.91mg/L) were associated with 1.4 times higher odds of 10-year incident cognitive impairment, after adjusting for race and age.^10^ Each of these three studies linked elevated cystatin C levels to worse cognitive status, and all concluded that the effect of cystatin C levels on cognitive status was homogeneous across racial/ethnic groups. Our analysis builds on these findings, by suggesting that the effect of cystatin C on dementia differs based on whether older adults are racialized as non-Hispanic Black or Hispanic rather than non-Hispanic White.

Of the previous studies, only the ABC study explored an interaction effect between race and cystatin C levels.^9^ Interaction effects are of public health importance because they illustrate which social groups are at most risk for adverse health events and could benefit from early interventions. However, statistical tests for interaction are scale dependent and test results may vary depending on the scale in which interaction effects are assessed—additive versus multiplicative.^39,40^ In the ABC study, researchers tested for a multiplicative interaction effect between race and cystatin C, concluding that no interaction was present.^9^ Nonetheless, epidemiologists prefer estimating interaction effects in the additive scale^41^ Our mediation-interaction approach allowed us to quantify two important components of the racial disparity: 1) the proportion due to the interaction between race/ethnicity and cystatin C (moderating pathway), and 2) the proportion due to the mediating effect of cystatin C.

We found that the effect of cystatin C on dementia may be modified by minoritized racial status, as evidenced by two of our measures: the excess risk due to interaction (a measure of additive interaction) and the proportion due to the interaction (a decomposition component of the mediation analysis). We observed that for minoritized participants with high levels of cystatin C (double exposed) the prevalence of dementia was higher than that of non-Hispanic White participants with low cystatin C (unexposed) or participants with either one of the two factors alone (single exposed). These findings held independently of demographic, behavioral, and other biomarkers. Mediation-interaction analysis results indicated that the interaction effect was responsible for explaining at least 9% percent of the racial disparity in prevalent dementia across both minoritized groups. The mediating effect of cystatin C only explained a small portion of the racial disparity for non-Hispanic Black participants and virtually none for Hispanic participants. The lack of mediation effect between Hispanic participants relative to non-Hispanic White participants may be because disparities in high levels of cystatin C across both groups are accounted for by covariates such as behavioral risk factors and other biomarkers, and because it is unlikely that cystatin C is the only pathway implicated in the production of racial disparities in prevalent dementia. Future research should focus on incorporating multiple mediators to better understand which deteriorated physiological systems are more likely to mediate racial disparities in cognitive aging.

Our sensitivity analyses demonstrated that our estimates were robust to the addition of *APOE-ε4* carrier status. *APOE-ε4* allele status is better understood as a precision variable rather than a confounder, given that its association is mostly established with dementia status and not cystatin C *per se*. Additionally, because dementia classification algorithms may have differential sensitivity and specificity performance across racial/ethnic groups, we also conducted our analyses using an alternative dementia classification method known to have similar sensitivity and specificity by racial groups.^34^ The use of Power’s algorithm for dementia classification dramatically attenuated the point estimates for our double exposed subgroups (non-Hispanic Black and Hispanic participants with high cystatin C) and hence their measures of additive interaction. Because the Power’s algorithm only classifies participants into two categories (dementia versus normal cognition), a sizable proportion of participants with cognitive impairment non-dementia (84%) or dementia (40.4%) according to the Langa-Weir algorithm may be classified as cognitively normal by the Power’s algorithm, which could attenuate any association between our variables of interest and dementia. In fact, our results found a super-additive effect between race/ethnicity and high cystatin C levels in participants with dementia but not with CIND which may help explain why our results using Power’s algorithm were indicative of no interaction in the additive scale.

This study’s strengths include quantification of the effect of cystatin C levels on two distinct cognitive outcomes in a large sample of non-Hispanic Black, Hispanic, and non-Hispanic White adults. We had rich data on several confounders, including a genetic risk factor for dementia and we conducted a sensitivity analysis to estimate the robustness of our results to potential misclassification of the outcome. Furthermore, we employed a novel statistical approach to accommodate effects due to interaction and mediation into a single framework. Limitations are that our data were cross-sectional, and thus subject to reverse causation and prevalence-incidence bias. Serum cystatin C levels may not be representative of cerebrospinal fluid concentrations, with limited direct relation to underlying neuropathological changes related to dementia.^12^ Our lack of repeated measures of cystatin C is an important limitation, and further studies should explore how changes in cystatin C levels may differ by race/ethnicity and differentially affect cognitive performance over time. Finally, our models did not adjust for glomerular filtration rate, and the depuration of circulating cystatin C may be affected by kidney functioning;^32,42^ therefore, we do not know to what extent our results may be susceptible to residual confounding effects. However, the ABC study showed that effect of cystatin C levels on cognition was independent of the effect of glomerular filtration rate.^9^

In conclusion, we observed that elevated serum cystatin C was associated with 1.4 times higher prevalence of dementia in a large, diverse sample of middle-aged and older US adults. The effect of cystatin C on prevalent dementia may be moderated by race/ethnicity, indicating that racialization affects not only the distribution of serum cystatin C across minoritized racial groups, but also the strength of association between the biomarker and dementia. Furthermore, we identified that non-Hispanic Black and Hispanic individuals with high levels of cystatin C are at increased risk for dementia. Public health efforts should focus on improving renal health outcomes in these vulnerable populations to ameliorate racial disparities in dementia. Future approaches should identify the role of other physiological pathways involved in the production of racial disparities in dementia. Our results provide compelling evidence that cystatin C is associated with adverse brain health, and this effect is larger than expected for individuals who are racialized as minorities. By implementing a novel methodological approach for causal mediation analysis, we illustrated how physiological biomarkers can be incorporated in the study of racial disparities in dementia.

## Data Availability

Survey data are publicly available (https://hrs.isr.umich.edu/data-products), and genetic data are available through dbGaP (https://dbgap.ncbi.nlm.nih.gov;phs000428.v2.p2).

https://github.com/bakulskilab

## Glossary

HRS: Health and Retirement Study,
PR: prevalence ratio,
CI: confidence interval,
TC: total cholesterol,
HDL-C: high density lipoprotein cholesterol,
HbA1C: glycosylated hemoglobin

## Acknowledgements

We thank the participants and staff of the Health and Retirement Study. We thank Brittni Delmaine for her excellent editorial services.

