## Supplemental Material for "Decomposing Interaction and Mediating Effects of Race/Ethnicity and Circulating Cystatin C on Cognitive Status in the United States Health and Retirement Study"

Additive interaction analysis

For our additive interaction analysis, we employed multivariable Poisson regression to estimate the joint effects of cystatin C and race/ethnicity on each of cognitive impairment non-dementia and dementia relative to normal cognition. We followed the same multivariable adjustment scheme described in the methods section and fit the additive interaction models for non-Hispanic Black and Hispanic participants, both relative to non-Hispanic White participants. We used regression coefficients to estimate three measures of interaction in the additive scale: the excess risk due to interaction, the attributable proportion, and the synergy index.[26] Positive values of these measures indicate departure from additivity and possible synergistic effect between the two interactive factors (race/ethnicity and high cystatin C). We used the delta method to calculate the standard errors and 95% CIs of the additive measures.

Mediation-interaction decomposition analysis

We conducted a mediation-interaction decomposition analysis using self-reported racialized categories as a proxy measure for exposure to racism and cystatin C as the mediator of the effect between racism and cognitive functioning. We employed a 4-way mediation and interaction decomposition analysis[22] to understand the overall effect of exposure to racism on cognitive impairment by identifying 1) the effect of the racialization process on cognitive status if the mediator (cystatin C) were set to concentrations $\leq$ 1.24mg/L. We refer to this effect as the "controlled direct effect" of racism (other pathways through which the racialization process operates besides cystatin C). We also identified 2) the reference interaction, which indicates the effect due to the interaction between race/ethnicity and cystatin C, setting the mediator to the levels of the most privileged racialized social group (non-Hispanic White); 3) the mediated interaction, or the interactive effect between race/ethnicity operating through the mediator (cystatin C), but allowing the levels of the mediator to vary across the compared racialized social groups; and 4) the pure indirect effect, or the effect of the mediator in the absence of the racialization process. From this decomposition, we estimated the attributable proportion due to each component, respective 95%CIs, and associated p-values. We calculated the decomposition effects in participants racialized as non-Hispanic Black and Hispanic separately and relative to participants racialized as non-Hispanic White.

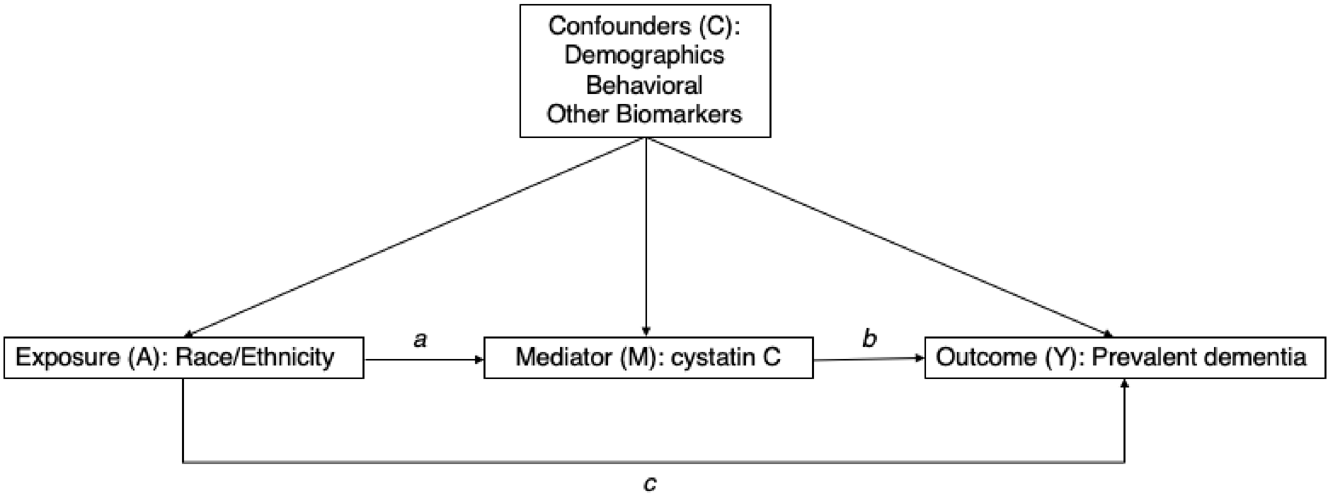


**Supplemental Figure 1:** Direct Acyclic Graph illustrating that the relationship between race/ethnicity (A) and prevalent dementia (Y) mediated through a marker of kidney health: cystatin C, denoted in the graph as M. **Note:** This mediation model accommodates interaction effects between race/ethnicity (A) and cystatin C (M), indicated through arrow *a*. Arrow *b* represents the effect of the mediator (M) on the outcome (Y). Arrow *c* illustrates the direct pathway from exposure (A) to outcome (Y).

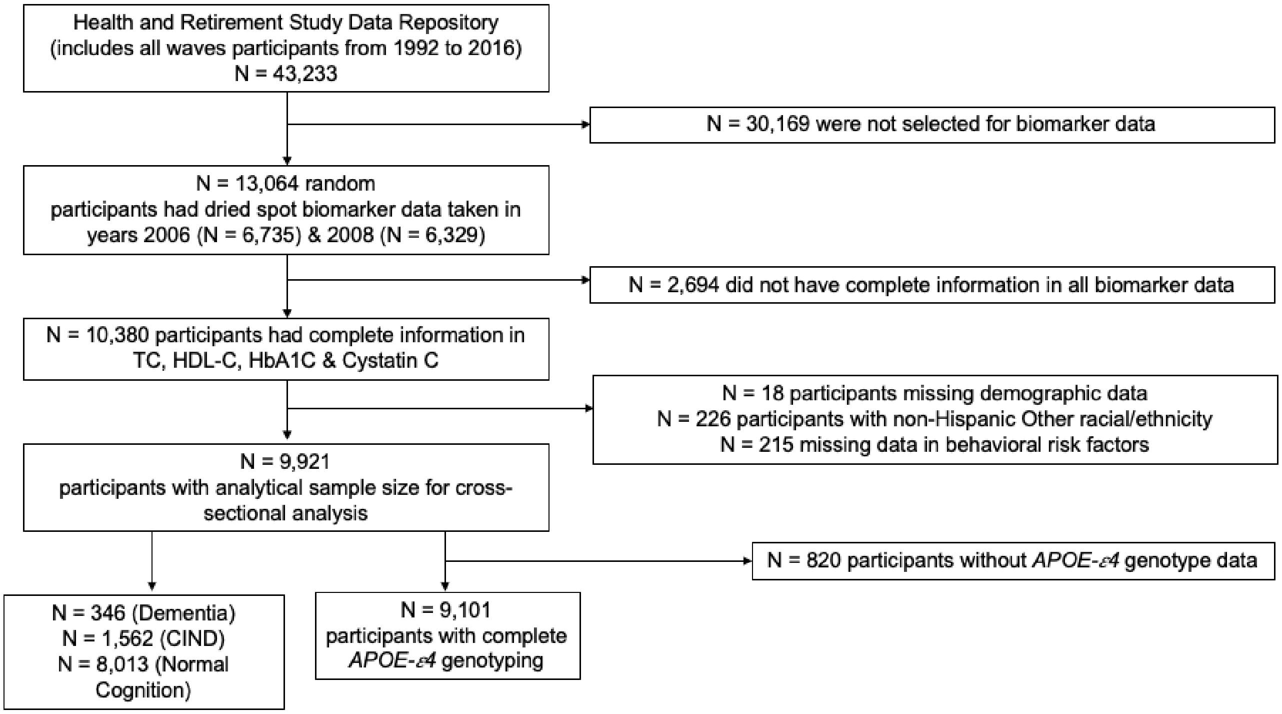


**Supplemental Figure 2:** Flow diagram of sample selection for cross-sectional study analysis of study participants in the Health and Retirement Study. CIND: cognitive impairment non-dementia; TC: total cholesterol; HDL-C: high density lipoprotein; HbA1C: glycosylated hemoglobin.

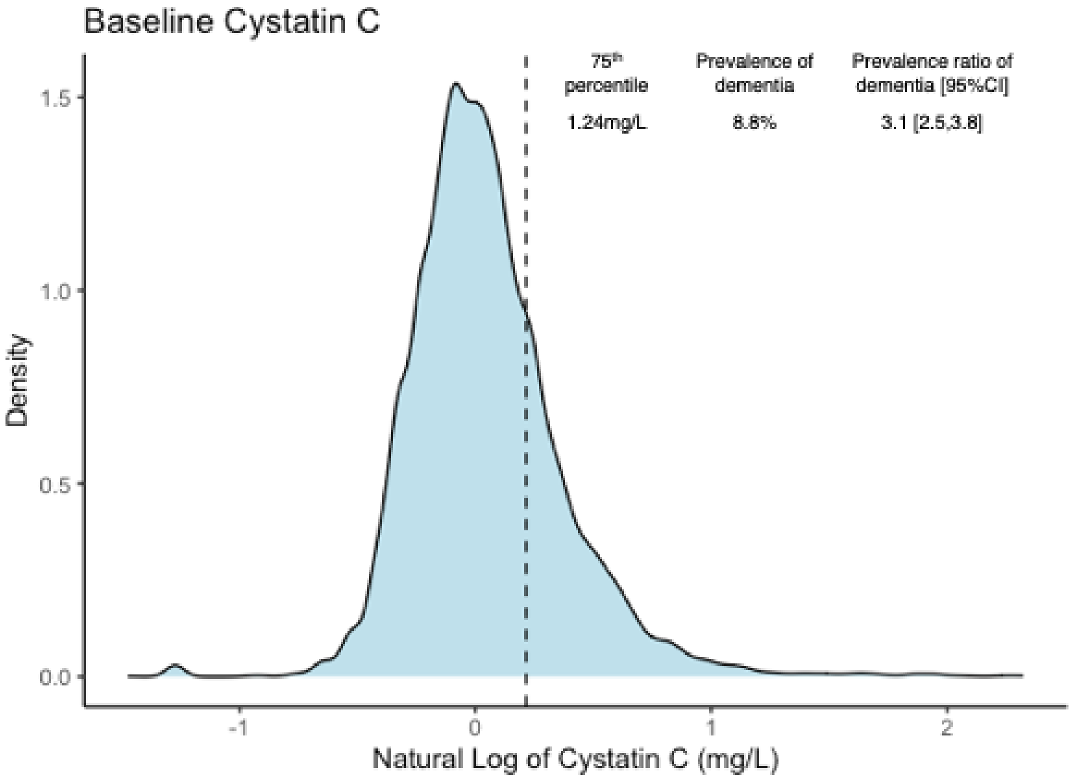


**Supplemental Figure 3:** Distribution of natural logarithm transformation of cystatin C using a Kernel density plot. The 75th percentile (or the log transformed value of cystatin C serum levels equal to 1.24mg/L) is indicated by the dotted line.

**Supplemental Table 1:** Distribution of baseline characteristics between included and excluded participants in the United States Health and Retirement Study, waves 2006 and 2008.

| | **Overall** | **Included** | **Excluded** | |
|---|---|---|---|---|
| Characteristic | N = 12,780[1] | N = 9,921[1] | N = 2,859[1] | p-value[2] |
| **Cognitive status** | | | | **<0.001** |
| Cognitively impaired non-dementia | 2,057 (16%) | 1,562 (16%) | 495 (17%) | |
| Dementia | 487 (3.8%) | 346 (3.5%) | 141 (4.9%) | |
| Normal | 10,236 (80%) | 8,013 (81%) | 2,223 (78%) | |
| **Race/ethnicity** | | | | 0.127 |
| non-Hispanic Black | 1,676 (13%) | 1,270 (13%) | 406 (14%) | |
| Hispanic | 1,188 (9.3%) | 918 (9.3%) | 270 (9.4%) | |
| non-Hispanic White | 9,916 (78%) | 7,733 (78%) | 2,183 (76%) | |
| **Gender** | | | | **0.016** |
| Female | 7,602 (59%) | 5,957 (60%) | 1,645 (58%) | |
| Male | 5,178 (41%) | 3,964 (40%) | 1,214 (42%) | |
| **Education Category** | | | | 0.099 |
| > College | 1,113 (8.7%) | 889 (9.0%) | 224 (7.8%) | |
| College/Some | 2,175 (17%) | 1,702 (17%) | 473 (17%) | |
| HS or < | 9,492 (74%) | 7,330 (74%) | 2,162 (76%) | |
| **Years of Education** | 12.53 (3.14) | 12.58 (3.12) | 12.38 (3.22) | **0.002** |
| **Age** (years) | 68.62 (10.54) | 68.37 (10.45) | 69.45 (10.79) | **<0.001** |
| **Alcohol** (# drinks/day when drinks) | 0.69 (1.40) | 0.69 (1.40) | 0.72 (1.42) | 0.359 |
| **Smoking** | | | | 0.789 |
| Current Smoker | 1,738 (14%) | 1,370 (14%) | 368 (13%) | |
| Former Smoker | 5,488 (43%) | 4,284 (43%) | 1,204 (44%) | |
| Never Smoker | 5,461 (43%) | 4,267 (43%) | 1,194 (43%) | |
| **Body Mass Index** (kg/$m^2$) | 28.21 (5.86) | 28.30 (5.86) | 27.90 (5.85) | **0.002** |
| **Hypertension** | | | | 0.569 |
| No | 5,639 (44%) | 4,369 (44%) | 1,270 (45%) | |
| Yes | 7,127 (56%) | 5,552 (56%) | 1,575 (55%) | |
| **Baseline Cystatin C** (mg/L) | 1.11 (0.53) | 1.12 (0.54) | 1.07 (0.49) | **<0.001** |
| **Baseline HbA1C** (%) | 5.87 (0.99) | 5.86 (0.97) | 5.93 (1.05) | **0.002** |
| **Baseline TC/HDL-C** (mg/dL) | 3.97 (1.20) | 3.97 (1.19) | 4.12 (1.33) | **0.009** |
| **Wave** | | | | **<0.001** |
| 2006 | 6,600 (52%) | 4,690 (47%) | 1,910 (67%) | |
| 2008 | 6,180 (48%) | 5,231 (53%) | 949 (33%) | |

[1] N (%); mean (SD)

[2] Pearson's chi-squared test; one-way ANOVA

HbA1C: glycosylated hemoglobin; TC/HDL-C: total cholesterol to high density lipoprotein-cholesterol ratio

**Supplemental Table 2:** Distribution of sample characteristics by the joint effect of race/ethnicity (non-Hispanic Black vs non-Hispanic White) and high cystatin C levels (>1.24mg/L) in the United States Health and Retirement Study, waves 2006 and 2008

| Characteristic | **Overall** N = 9,003[1] | **non-Hispanic White & Low cystatin C[*] (Unexposed)** N = 5,902[1] | **Non-Hispanic White & High cystatin C[*] (Single exposed)** N = 1,831[1] | **non-Hispanic Black & Low cystatin C[*] (Single exposed)** N = 932[1] | **non-Hispanic Black & High cystatin C[*] (Double Exposed)** N = 338[1] | p-value[2] |
|---|---|---|---|---|---|---|
| **Dementia status** | | | | | | **<0.001** |
| Cognitively normal | 7,395 (96%) | 5,229 (98%) | 1,395 (95%) | 624 (92%) | 147 (74%) | |
| Dementia | 288 (3.7%) | 100 (1.9%) | 81 (5.5%) | 56 (8.2%) | 51 (26%) | |
| **Impairment status** | | | | | | **<0.001** |
| Cognitively normal | 7,395 (85%) | 5,229 (90%) | 1,395 (80%) | 624 (71%) | 147 (51%) | |
| Cognitive impairment non-dementia | 1,320 (15%) | 573 (9.9%) | 355 (20%) | 252 (29%) | 140 (49%) | |
| **Gender** | | | | | | **<0.001** |
| Female | 5,371 (60%) | 3,478 (59%) | 1,061 (58%) | 614 (66%) | 218 (64%) | |
| Male | 3,632 (40%) | 2,424 (41%) | 770 (42%) | 318 (34%) | 120 (36%) | |
| **Education** | | | | | | **<0.001** |
| > College | 868 (9.6%) | 676 (11%) | 128 (7.0%) | 50 (5.4%) | 14 (4.1%) | |
| College/Some | 1,630 (18%) | 1,212 (21%) | 269 (15%) | 123 (13%) | 26 (7.7%) | |
| High school or < | 6,505 (72%) | 4,014 (68%) | 1,434 (78%) | 759 (81%) | 298 (88%) | |
| **Age** (years) | 68.70 (10.39) | 66.98 (9.82) | 75.28 (9.88) | 65.44 (9.25) | 72.26 (9.97) | **<0.001** |
| **Years of Education** | 12.93 (2.70) | 13.31 (2.52) | 12.58 (2.60) | 11.98 (3.00) | 10.87 (3.39) | **<0.001** |
| **Alcohol** (# drinks/day when drinks) | 0.70 (1.32) | 0.82 (1.40) | 0.43 (1.01) | 0.57 (1.39) | 0.30 (0.88) | **<0.001** |
| **Smoking Status** | | | | | | **<0.001** |
| Current | 1,255 (14%) | 793 (13%) | 235 (13%) | 183 (20%) | 44 (13%) | |
| Former | 3,927 (44%) | 2,574 (44%) | 849 (46%) | 352 (38%) | 152 (45%) | |
| Never | 3,821 (42%) | 2,535 (43%) | 747 (41%) | 397 (43%) | 142 (42%) | |
| **Body Mass Index** (kg/m$^2$) | 28.20 (5.87) | 27.63 (5.40) | 28.76 (6.42) | 29.92 (6.32) | 30.47 (7.44) | **<0.001** |

**Supplemental Table 2:** Distribution of sample characteristics by the joint effect of race/ethnicity (non-Hispanic Black vs non-Hispanic White) and high cystatin C levels (>1.24mg/L) in the United States Health and Retirement Study, waves 2006 and 2008

| Characteristic | **Overall** N = 9,003[1] | **non-Hispanic White & Low cystatin C[*] (Unexposed)** N = 5,902[1] | **Non-Hispanic White & High cystatin C[*] (Single exposed)** N = 1,831[1] | **non-Hispanic Black & Low cystatin C[*] (Single exposed)** N = 932[1] | **non-Hispanic Black & High cystatin C[*] (Double Exposed)** N = 338[1] | p-value[2] |
|---|---|---|---|---|---|---|
| **Hypertension** | | | | | | **<0.001** |
| No | 3,947 (44%) | 3,139 (53%) | 467 (26%) | 298 (32%) | 43 (13%) | |
| Yes | 5,056 (56%) | 2,763 (47%) | 1,364 (74%) | 634 (68%) | 295 (87%) | |
| **Baseline HbA1C** (%) | 5.82 (0.91) | 5.70 (0.80) | 5.94 (0.91) | 6.19 (1.24) | 6.27 (1.18) | **<0.001** |
| **Baseline TC/HDL-C** (mg/dL) | 3.95 (1.19) | 3.93 (1.18) | 4.08 (1.22) | 3.83 (1.14) | 3.89 (1.15) | **<0.001** |

[1]Statistics presented: N (Col%); Mean (SD)

[2]Statistical tests performed: chi-square test of independence; One-way ANOVA

[*]High cystatin C defined as serum levels > 1.24mg/L (> 75th percentile of sample distribution) and low cystatin C defined as ≤1.24mg/L

TC/HDL-C: total cholesterol to high density lipoprotein-cholesterol ratio; HbA1C: glycosylated hemoglobin

**Supplemental Table 3:** Distribution of sample characteristics by the joint effect of ethnicity (Hispanic vs non-Hispanic White) and high cystatin C levels (>1.24mg/L) in the United States Health and Retirement Study, waves 2006 and 2008

| Characteristic | **Overall** N = 8,651[1] | **non-Hispanic White & Low cystatin C*** **(Unexposed)** N = 5,902[1] | **non-Hispanic White & High cystatin C*** **(Single Exposed)** N = 1,831[1] | **Hispanic & Low cystatin C*** **(Single exposed)** N = 753[1] | **Hispanic & High cystatin C*** **(Double exposed)** N = 165[1] | p-value[2] |
|---|---|---|---|---|---|---|
| **Dementia status** | | | | | | **<0.001** |
| Cognitively normal | 7,242 (97%) | 5,229 (98%) | 1,395 (95%) | 530 (94%) | 88 (78%) | |
| Dementia | 239 (3.2%) | 100 (1.9%) | 81 (5.5%) | 33 (5.9%) | 25 (22%) | |
| **Cognitively impaired non-dementia** | | | | | | **<0.001** |
| Cognitively normal | 7,242 (86%) | 5,229 (90%) | 1,395 (80%) | 530 (74%) | 88 (63%) | |
| Cognitively impaired non-dementia | 1,170 (14%) | 573 (9.9%) | 355 (20%) | 190 (26%) | 52 (37%) | |
| **Gender** | | | | | | **0.018** |
| Female | 5,125 (59%) | 3,478 (59%) | 1,061 (58%) | 483 (64%) | 103 (62%) | |
| Male | 3,526 (41%) | 2,424 (41%) | 770 (42%) | 270 (36%) | 62 (38%) | |
| **Education** | | | | | | **<0.001** |
| > College | 825 (9.5%) | 676 (11%) | 128 (7.0%) | 18 (2.4%) | 3 (1.8%) | |
| College/Some | 1,553 (18%) | 1,212 (21%) | 269 (15%) | 63 (8.4%) | 9 (5.5%) | |
| High school or < | 6,273 (73%) | 4,014 (68%) | 1,434 (78%) | 672 (89%) | 153 (93%) | |
| **Age** (years) | 68.54 (10.52) | 66.98 (9.82) | 75.28 (9.88) | 63.41 (9.82) | 73.07 (9.80) | **<0.001** |
| **Years of Education** | 12.71 (3.09) | 13.31 (2.52) | 12.58 (2.60) | 9.35 (4.52) | 8.21 (4.44) | **<0.001** |
| **Alcohol** (# drinks/day when drinks) | 0.72 (1.41) | 0.82 (1.40) | 0.43 (1.01) | 0.62 (1.40) | 0.55 (3.62) | **<0.001** |
| **Smoking Status** | | | | | | **<0.001** |
| Current | 1,143 (13%) | 793 (13%) | 235 (13%) | 105 (14%) | 10 (6.1%) | |
| Former | 3,780 (44%) | 2,574 (44%) | 849 (46%) | 285 (38%) | 72 (44%) | |
| Never | 3,728 (43%) | 2,535 (43%) | 747 (41%) | 363 (48%) | 83 (50%) | |

**Supplemental Table 3:** Distribution of sample characteristics by the joint effect of ethnicity (Hispanic vs non-Hispanic White) and high cystatin C levels (>1.24mg/L) in the United States Health and Retirement Study, waves 2006 and 2008

| Characteristic | **Overall** N = 8,651[1] | **non-Hispanic White & Low cystatin C*** **(Unexposed)** N = 5,902[1] | **non-Hispanic White & High cystatin C*** **(Single Exposed)** N = 1,831[1] | **Hispanic & Low cystatin C*** **(Single exposed)** N = 753[1] | **Hispanic & High cystatin C*** **(Double exposed)** N = 165[1] | p-value[2] |
|---|---|---|---|---|---|---|
| **Body Mass Index** (kg/m$^2$) | 28.04 (5.69) | 27.63 (5.40) | 28.76 (6.42) | 29.05 (5.42) | 30.06 (6.58) | **<0.001** |
| **Hypertension** | | | | | | **<0.001** |
| No | 4,028 (47%) | 3,139 (53%) | 467 (26%) | 385 (51%) | 37 (22%) | |
| Yes | 4,623 (53%) | 2,763 (47%) | 1,364 (74%) | 368 (49%) | 128 (78%) | |
| **Baseline HbA1C** (%) | 5.81 (0.92) | 5.70 (0.80) | 5.94 (0.91) | 6.18 (1.41) | 6.47 (1.42) | **<0.001** |
| **Baseline TC/HDL-C** (mg/dL) | 3.98 (1.20) | 3.93 (1.18) | 4.08 (1.22) | 4.15 (1.26) | 3.99 (1.13) | **<0.001** |

[1]Statistics presented: N (Col%); Mean (SD)

[2]Statistical tests performed: chi-square test of independence; One-way ANOVA

*High cystatin C defined as serum levels > 1.24mg/L (> 75$^{th}$ percentile of sample distribution) and low cystatin C defined as ≤1.24mg/L

TC/HDL-C: total cholesterol to high density lipoprotein-cholesterol ratio; HbA1C: glycosylated hemoglobin

**Supplemental Table 4**: Prevalence ratios and measures of interaction in the additive scale of the joint effect of race/ethnicity and high cystatin C (>1.24mg/L) on dementia in the United States health and Retirement Study, waves 2006 & 2008

| | **Demographic Model and *APOE-ε4* adjustment** | | | |
|---|---|---|---|---|
| | **Non-Hispanic Black[+]** | **95%CI** | **Hispanic[+]** | **95%CI** |
| **Dementia** | | | | |
| Unexposed[a] | 1 | - | 1 | - |
| Single exposed[b] (CysC) | 1.3 | [0.9,1.7] | 1.2 | [0.9,1.7] |
| Single exposed[c] (minority) | 4.9*** | [3.6,6.8] | 4.2*** | [2.9,6.1] |
| Double exposed[d] | 7.2*** | [5.3,9.8] | 5.9*** | [3.8,9.2] |
| | **Measures of Interaction in the Additive Scale** | | | |
| Excess risk due to interaction[†] | 2.0 | [-0.1,4.1] | 1.5 | [-1.2,4.2] |
| Attributable proportion[‡] | 0.3 | [0.0, 0.5] | 0.3 | [-0.1,0.6] |
| Synergy index[£] | 1.5 | [1.0, 2.2] | 1.4 | [0.7,2.7] |
| **Observations** | 7,051 | | 6,886 | |

Prevalence ratios; 95% confidence intervals in brackets

* $p < 0.05$, ** $p < 0.01$, *** $p < 0.001$

[+] Model adjusting for demographic variables (age, gender, education) and frequency of any *APOE-ε4* allele

a) Unexposed group (reference group): non-Hispanic White & low cystatin C (≤1.24mg/L)

b) Single exposed (CysC): non-Hispanic White & high cystatin C (>1.24mg/L)

c) Single exposed (minority): either non-Hispanic Black or Hispanic & low cystatin C (≤1.24mg/L)

d) Double exposed: either non-Hispanic Black or Hispanic & high cystatin C (>1.24mg/L)

† Excess risk = Prevalence ratio (PR) of double exposed - PR of single exposed (minority) - PR of single exposed (CysC) + 1

‡ Attributable proportion = [Excess risk due to interaction] / [PR of double exposed]

£ Synergy index = [PR of double exposed - 1] / [PR of single exposed (minority) + PR of single exposed (CysC) - 2]

**Supplemental Table 5A:** Dementia model exploring the joint effect of race (non-Hispanic Black) and high serum levels of cystatin C (>1.24mg/L) in the United States Health and Retirement Study using the Power's algorithm for outcome classification

| | Unadjusted | | Demographic[1] | | Behavioral[2] | | Biomarker[3] | |
|---|---|---|---|---|---|---|---|---|
| **Dementia** | | | | | | | | |
| Unexposed[a] | 1 | - | 1 | - | 1 | - | 1 | - |
| Single exposed[b] (CysC) | 3.9*** | [3.2,4.7] | 1.3* | [1.0,1.5] | 1.3* | [1.0,1.5] | 1.2* | [1.0,1.5] |
| Single exposed[c] (minority) | 1.5* | [1.1,2.1] | 1.9*** | [1.4,2.6] | 1.9*** | [1.4,2.7] | 1.9*** | [1.4,2.6] |
| Double exposed[d] | 6.0*** | [4.5,7.9] | 2.6*** | [2.0,3.4] | 2.6*** | [2.0,3.4] | 2.6*** | [2.0,3.4] |
| **Observations** | 8,951 | | 8,951 | | 8,951 | | 8,951 | |

Prevalence ratios; 95% confidence intervals in brackets

* $p < 0.05$, ** $p < 0.01$, *** $p < 0.001$

[1] Demographic model: adjusted for age, sex, education

[2] Behavioral model: adjusted for age, sex, education, smoking status, alcohol consumption, body mass index

[3] Biomarker model: adjusted for age, sex, education, smoking status, alcohol consumption, body mass index, total cholesterol to high density lipoprotein-cholesterol ratio, glycosylated hemoglobin, and hypertension

[a)] Unexposed group (reference group): non-Hispanic White & low cystatin C (≤1.24mg/L)

[b)] Single exposed (CysC): non-Hispanic White & high cystatin C (>1.24mg/L)

[c)] Single exposed (minority): non-Hispanic Black & low cystatin C (≤1.24mg/L)

[d)] Double exposed: non-Hispanic Black & high cystatin C (>1.24mg/L)

**Supplemental Table 5B:** Dementia model exploring the joint effect of ethnicity (Hispanic) and high serum levels of cystatin C (>1.24mg/L) in the United States Health and Retirement Study using the Power's algorithm for outcome classification

| | Unadjusted | | Demographic[1] | | Behavioral[2] | | Biomarker[3] | |
|---|---|---|---|---|---|---|---|---|
| **Dementia** | | | | | | | | |
| Unexposed[a] | 1 | - | 1 | - | 1 | - | 1 | - |
| Single exposed[b] (CysC) | 3.9*** | [3.2,4.7] | 1.2* | [1.0,1.5] | 1.2 | [1.0,1.5] | 1.2 | [0.9,1.4] |
| Single exposed[c] (minority) | 0.7 | [0.4,1.1] | 1.0 | [0.6,1.7] | 1.0 | [0.6,1.6] | 0.9 | [0.5,1.5] |
| Double exposed[d] | 3.5*** | [2.1,5.7] | 1.4 | [0.9,2.1] | 1.3 | [0.9,2.0] | 1.3 | [0.8,1.9] |
| **Observations** | 8,602 | | 8,602 | | 8,602 | | 8,602 | |

Prevalent ratios; 95% confidence intervals in brackets

* $p < 0.05$, ** $p < 0.01$, *** $p < 0.001$

[1] Demographic model: adjusted for age, sex, education

[2] Behavioral model: adjusted for age, sex, education, smoking status, alcohol consumption, body mass index

[3] Biomarker model: adjusted for age, sex, education, smoking status, alcohol consumption, body mass index, total cholesterol to high density lipoprotein-cholesterol ratio, glycosylated hemoglobin, and hypertension

a) Unexposed group (reference group): non-Hispanic White & low cystatin C (≤1.24mg/L)

b) Single exposed (CysC): non-Hispanic White & high cystatin C (>1.24mg/L)

c) Single exposed (minority): Hispanic & low cystatin C (≤1.24mg/L)

d) Double exposed: Hispanic & high cystatin C (>1.24mg/L)

**Supplemental Table 6**: Prevalence ratios and measures of interaction in the additive scale of the joint effect of race/ethnicity and high cystatin C (>1.24mg/L) on dementia in the United States health and Retirement Study using the Power's algorithm for dementia classification, waves 2006 & 2008

| | Demographic Model | | | |
|---|---|---|---|---|
| | **Non-Hispanic Black+** | **95%CI** | **Hispanic+** | **95%CI** |
| **Dementia** | | | | |
| Unexposed[a] | 1 | - | 1 | - |
| Single exposed[b] (CysC) | 1.3* | [1.0,1.5] | 1.2* | [1.0,1.5] |
| Single exposed[c] (minority) | 1.9*** | [1.4,2.6] | 1.0 | [0.6,1.7] |
| Double exposed[d] | 2.6*** | [2.0,3.4] | 1.4 | [0.9,2.1] |
| | **Measures of Interaction in the Additive Scale** | | | |
| Excess risk due to interaction† | 0.4 | [-0.4, 1.2] | 0.1 | [-0.7, 0.9] |
| Attributable proportion‡ | 0.2 | [-0.1, 0.5] | 0.1 | [-0.5, 0.6] |
| Synergy index£ | 1.4 | [0.7, 2.5] | 1.4 | [-0.1, 16.4] |
| **Observations** | 8,951 | | 8,602 | |

Prevalence ratios; 95% confidence intervals in brackets

* $p < 0.05$, ** $p < 0.01$, *** $p < 0.001$

+ Model adjusting for demographic variables (age, gender, education)

a) Unexposed group (reference group): non-Hispanic White & low cystatin C (≤1.24mg/L)

b) Single exposed (CysC): non-Hispanic White & high cystatin C (>1.24mg/L)

c) Single exposed (minority): either non-Hispanic Black or Hispanic & low cystatin C (≤1.24mg/L)

d) Double exposed: either non-Hispanic Black or Hispanic & high cystatin C (>1.24mg/L)

† Excess risk = Prevalence ratio (PR) of double exposed - PR of single exposed (minority) - PR of single exposed (CysC) + 1

‡ Attributable proportion = [Excess risk due to interaction] / [PR of double exposed]

£ Synergy index = [PR of double exposed - 1] / [PR of single exposed (minority) + PR of single exposed (CysC) - 2
